# When the unexpected pandemic began, what were the experiences of healthcare professionals in the management of COVID-19 in Lesotho?

**DOI:** 10.1101/2025.08.01.25332516

**Authors:** Isabel Nyangu, Maseabata Ramathebane

**Affiliations:** School of Health and Social Care, Edinburgh Napier University; Department of Pharmacy, National University of Lesotho

**Keywords:** COVID-19, healthcare professionals, experiences, Lesotho, pandemic

## Abstract

In Lesotho, the healthcare system was not prepared to deal with the COVID-19 pandemic. While PPE availability was a problem throughout the world, for resource-limited countries like Lesotho, the problem was even bigger. Additionally, community transmission became a problem because there was a backlog of tests, delaying the results of COVID-19 tests.

This study aimed to explore and describe the experiences of HCPs on the management of the COVID-19 pandemic in Lesotho. An explorative descriptive qualitative design was used to collect data from healthcare professionals who were purposively sampled from five health facilities and took part in focus group discussions. Written informed consent was sought from the participants who voluntarily took part in the study. They were allowed to ask questions and could withdraw from the study without any repercussions. Constant comparison analysis was used to analyse data and was presented using themes, subthemes, and categories.

COVID-19 caused unpleasant emotional effects among healthcare professionals as they feared becoming infected and were faced with stigma and discrimination in their own families. They had limited protection from COVID-19 as there was inadequate personal protective equipment, and they could not effectively separate patients who came to the facilities. The support they received varied and was not adequate to address their needs during the pandemic. They lacked information about the pandemic, did not receive any psychological support, and they felt that more assistance could have helped them better manage the pandemic.

It remains crucial for healthcare professionals to be empowered with the correct and accurate information, personal protective equipment, and psychological support when faced with pandemics like COVID-19.

## Introduction

As a country, Lesotho had to take serious measures in its Covid-19 response, particularly because the population is already faced with alarming rates of HIV and TB, and increasing non-communicable diseases such as heart diseases, diabetes mellitus, hypertension, and cancers which are predisposing diseases to high morbidity and mortality rates of Covid-19 (LDHS, 2016; Sengupta et al., 2021). The health system was not prepared to deal with the pandemic, and due to its rapid spread, HCPs were at risk of contracting COVID-19 and passing it to their family members (Bandyopadhyay et al., 2020). The need for personal protective equipment (PPE) was insurmountable. While PPE availability was a problem throughout the world, for resource-limited countries like Lesotho, the problem was even bigger. Again, a general lack of healthcare infrastructure and standard operating procedures put the lives of HCPs at risk of contracting the virus, worsening the performance of the already overstretched and poor health system (Khalid & Ali, 2020; Desouky et al., 2021).

The study of Tan et al. (2020) raised an important matter of non-frontline HCPs who had less information regarding the importance of PPE and infection control measures, leading to less support for frontline workers. It is important for medical information on COVID-19 to come from trusted sources, even for non-frontline HCPs (Norhayati et al., 2021). Social media played a major role in circulating unverified information about the number of patients and the death toll, which led HCPs to suffer from depression (Aghili & Arbabi, 2020).

The fact that most patients with COVID-19 are asymptomatic puts healthcare professionals at risk of being infected, and the problem is worsened by delayed results due to limited testing capacity (Bandyopadhyay et al., 2020). This further means that if HCPs become infected and asymptomatic, they infect both patients and family members, particularly if their knowledge about the risks is limited and may lead to erroneous practices (Polychronis & Roupa, 2020).

In Lesotho, community transmission became a problem because there was a backlog of tests, delaying the results of COVID-19 tests. Similarly, Ogbolosingha & Singh (2020) believed widespread community transmission in several countries in sub-Saharan Africa was due to the delayed detection of COVID-19. It was further advocated that all symptomatic individuals and their contacts should be tested for COVID-19 (Ogbolosingha & Singh, 2020). Another disturbing fact is the mental well-being of HCPs in resource-limited countries, as Carrieri et al. (2018) indicated that the mental health and psychological well-being of HCPs working under enormous pressure was already identified as a major healthcare issue. There was the growing incidence of stress, burnout, depression, drug and alcohol dependence and suicide across all groups of HCPs, in many countries (Carrieri et al., 2018; Norhayati et al., 2021). What is important is that this was identified long before the current COVID-19 pandemic (Carrieri et al., 2018; Tan et al., 2020). Again, Shaukat et al. (2020) connected the increased psychological impacts with high exposure risk due to a lack of PPE and long working hours.

### Aim

This study aimed to explore and describe the experiences of HCPs on the management of the COVID-19 pandemic in Lesotho.

## Methodology

A descriptive qualitative design was used to collect data from 14 healthcare professionals who were purposively sampled. They included nurse assistants, registered nurse midwives, nurse clinicians, and pharmacists who were working at five health centres. A pre-test of the focus group guide was conducted to ascertain its trustworthiness. Data were collected from September 2021 until November 2021 by trained data collectors. Audio tapes were used to record the discussions, and data were analysed using constant comparison analysis and is presented using themes, sub-themes, and categories.

### Ethical Considerations

The study and all its procedures were submitted to the National University of Lesotho (NUL) Institutional Research Board (IRB), the Lesotho Ministry of Health Research and Ethics Committee (ID20-2021). Respondents were asked to give written informed consent to take part in the study. A confidentiality agreement was signed to ensure privacy. Only codes were used to identify respondents, and all research data was encrypted and stored in a lockable cupboard. Virtues of sensitivity, respect and patience were applied during contact periods with respondents who were asked to voluntarily take part in the study and could withdraw without any prejudice. Aspects of the study were fully disclosed to the participants, including the likely risks and benefits.

## Results

A total of 14 healthcare professionals took part in the focus group discussions. Table 1 below shows the distribution of participants who took part in the focus group discussions.

**Table 1:** Participants.

| Cadre | Number |
| --- | --- |
| Nurse clinicians | 1 |
| Registered nurse midwives | 9 |
| Nurse assistants | 1 |
| Pharmacists | 3 |

### Theme 1: COVID-19 caused unpleasant emotional effects

This theme describes how COVID-19 affected the respondents emotionally. It has three sub-themes of devastation, terror, stigma and discrimination.

### Sub-theme 1: Emotional Devastation

Respondents described how they were distraught due to COVID-19, feeling shocked, traumatised, and confused.

> *Covid-19 was not expected; it was scary; We were scared as we had to face those with signs of flu; We did not know how to react* (pharmacists)

Healthcare workers were not prepared for the pandemic at all, which made them even more frightened.

> *I think it was scary for me because we were not prepared; The experience was not a good one*; *I was panicking; I was traumatised and wanted to leave the profession. (nurses)*

### Sub-theme 2: Terror due to fear of contagion

Respondents described how they were frightened of COVID-19 as they underwent a fear of the unknown and anticipated death.

> *We just knew it was contagious: you got infected by just touching someone* (nurses)
>
> *We had the fear of being infected; We also feared death as many people were dying, including health care workers*. (pharmacists)

### Sub-theme 3: Stigma and Discrimination

Respondents also experienced dishonour and prejudice due to COVID-19 amongst themselves and in their own families.

> *We had to be isolated from our families: they were afraid of us as we could carry the virus to them* (nurses)
>
> They also feared infecting their loved ones with the disease.
>
> *We feared contracting the disease and then taking it home to the family*. (nurses)

### Theme 2: Healthcare workers had limited protection from COVID-19

This theme describes how health care professionals struggled to obtain personal protective equipment and could not isolate patients adequately during the COVID-19 pandemic.

### Sub-theme 1 Inadequate personal protective equipment (PPE)

Respondents explained how they did not have adequate protective equipment to assist patients who presented with COVID-19.

> *We did not have any PPE from work, and we were wearing our own usual nurses’ uniform clothing’* (nurses)
>
> Student nurses did not have access to PPE whilst on clinical placement.
>
> *When the pandemic started, we were still students and were not allowed to use any of the PPE at the facility*. (nurses)

There seemed to be misunderstanding as to who was entitled to receive PPE amongst healthcare staff.

> *Initially we were sidelined from being given PPE as compared to doctors and nurses and yet the same patients they saw also had to come and collect medications from the pharmacy; initially we were not given PPE, we did not get even surgical masks; But eventually we got it*. (pharmacists)

Equipment that was crucial in the management of patients was also not available.

> *We did not have any intensive care unit equipment*.

### Sub-theme 2 Inability to isolate patients adequately

Respondents described how difficult it was to separate patients who came seeking health care services due the existing healthcare facility infrastructure.

> *The clinic settings do not allow for isolation of patients as they do not reveal their true symptoms during screening*. (nurses)

This was a challenge even when medications were dispensed to patients.

> *Since all patients collect medications from us, we had to give medications outside using a trolley, which was not ethical in terms of pharmacy practice. Some patients understood, whilst others did not*. (pharmacists)

### Theme 3: Varying Support was given to health care workers

This theme explains the different types of support (if any) that were availed to health care workers at their facilities during the pandemic.

### Sub-theme 1 There was some effort from employers to support staff

Respondents explained how they were given ample support from their employers through education and PPE supplies.

> *They started by educating us and then providing PPE. They were very much supportive* (nurses)

Even partner organisations to the government assisted the facilities.

> *PPE from the non-governmental organisations were received* (nurses)

### Sub-theme 2 Support provided was not enough

Whilst the support that was provided was appreciated, respondents also explained how the support they were given was not adequate, as they could not give adequate health education to patients, the PPE was limited, and the risk allowance was not always accessible for all.

> *Support was not adequate* (nurses and pharmacists)

Nurses felt they could not provide enough information on the pandemic, as they knew little about it as well.

> *We could not even provide information adequately as we did not have the knowledge ourselves* (nurses)

The protective equipment was also not enough for all healthcare workers.

> *PPE was not adequate* (pharmacists)

Financial support for risk allowance was not accessed by all healthcare workers.

> *We only get risk allowance occasionally, and it is not always available, but the new COVID-19 cases always exist*. (nurses)

### Sub-theme 3 No psychological support was provided

Respondents explained how there was no psychological support during the COVID-19 pandemic. Nurses felt that there was no one to assist with their mental health.

> *There was no one to soothe your psychological well-being* (nurses) Healthcare workers needed psychotherapy to assist them during the pandemic.

> *We needed counselling, but we did not get it. We had to sort our mindsets alone* (pharmacists)

### Sub-theme 4 Limited education given to healthcare professionals

Information on COVID-19 was also very limited, and the education provided to healthcare workers was not adequate.

> *We did not know about it; there was no information; Covid-19 arrived in a rush; we had little training* (nurses)

### Theme 4 More assistance was necessary to improve service provision during the pandemic

This theme describes suggestions for improving the health care service delivery during the COVID-19 pandemic. Respondents suggested more training and information on COVID-19, more human resources, and better infrastructure.

Healthcare workers proposed more intensive training so they could understand the pandemic.

> *We mostly need training at healthcare facilities, especially on new issues on the disease* (nurses).
>
> *More sensitisation of staff is necessary for us to prepare adequately, including giving us knowledge and involving us more in decision making* (pharmacists)

It was suggested that more healthcare staff were needed to ensure that all people are served.

> *We need more staff so that there are those dedicated to those suspected of having Covid-19; This will ensure that other patients presenting with other conditions are cared for*. (nurses)
>
> A provision for COVID-19 isolation had to be considered to reduce the spread of the pandemic.
>
> *‘We have a problem of infrastructure; the buildings are packed and cannot cater for those with Covid-19; it makes other patients uncomfortable, hence we need better isolation facilities*.*’* (nurses)

Social media platforms had to be considered to improve communication in the public.

‘W*e can have radio programs to improve public knowledge on Covid-19’*. (nurses)

## Discussion

The study brings a better understanding of the experiences of HCPs in Lesotho, which is like the experience of HCPs all over the world. According to the study of Shahbaz et al. (2021) indicated that HCPs had challenges lack of psychosocial support, during post-duty quarantine. This study reported that sharing information about COVID-19 was lacking among hospitalised HCPs, yet HCPs are supposed to provide accurate and relevant information to the patients. However, the study of Ogbolosingha and Singh (2020) stated that for successful repression of the number of COVID-19 cases, it is important to provide health education and public sensitisation about the pandemic.

According to Shaban et al. (2020), isolation of patients was done to limit the COVID-19 infection. However, the physical characteristics of the environment in which patients are isolated may affect their experiences (Barratt et al., 2011; Abad et al., 2010). HCPs advised that Covid-19 education is critical, and this is also mentioned by Billings et al. (2021), who stipulated that HCPs valued clear, consistent, and compassionate communication.

Commonly, the highest number of HCPs feared contracting Covid-19, followed by those who feared death, and those who feared infecting their families. Equally, Yin and Zeng highlighted the need for psychological support for HCPs during the COVID-19 pandemic to ease tensions and fears. Again, the results show that HCPs feared infecting family members, as most of them still stayed at home during the pandemic, Sengupta et al. (2021) indicated that nurses feared infecting family members and had to sanitise everything, including their cellular phone, before they could interact with their children.

The support given was not adequate, such that they too could not educate the patients appropriately, the PPE was not adequate, and the risk allowance was not always accessible for all. Conversely, according to Billings et al. 2021, HCPs feel valued when their safety and support in terms of workloads and time out of work are considered a priority and are provided by their organisations. Some HCPs indicated that patients with COVID-19 die, so it makes them sad. This is not new, as it was clearly stated by Bayuo and Agbenorku, (2017); Kisorio and Langley (2019) that HCPs experience emotional and physical exhaustion when they care for critically ill patients.

However, those who were involved in the focus group discussion felt isolated by the communities in which they lived. The study of Billings et al. (2021) stated that HCPS sometimes considered relationships with families, colleagues, organisations, the media and the wider public to be complicated and associated them with either support or sources of stress.

## Limitations of the study

Data collection was done at the same time as MOH was rolling out community COVID-19 vaccination, and this caused a delay in permission to carry out the study at the districts.

## Conclusion

The conclusion that can be drawn regarding the views about experiences of HCPs is that their personal experience was filled with fear, shock and anxiety due to the high infection and mortality rate, which was perpetuated by shortage of PPE, lack of infrastructure, shortage of staff, and lack of information. Limited information about the disease also played a major part, as HCPs were supposed to educate patients when they were lacking in knowledge. All these led to compromised service provision to both COVID-19-infected patients and other patients who came to the facilities with other general conditions.

## Recommendations

- HCPs expressed the need to be empowered with correct and accurate information about COVID-19 as they pass this information to patients.
- The fear of contracting COVID-19 was propagated by a lack of enough PPE; therefore, it is important to make sure that there is enough PPE.
- The risk allowances must be given to all healthcare staff members who come in contact with patients infected with COVID-19.
- Infrastructure at the health facilities must be improved so that the coming waves of COVID-19 are better managed and allow for the isolation of patients. For example, temporary shelters can be used.
- It is paramount to provide psychosocial and mental well-being support to all HCPs.

## Data Availability

All data produced in the present work are contained in the manuscript.

